# *Hanging on through Omicron, then what*? A pre-exit baseline of the U.S. emergency nursing workforce, 2018–2022, with implications for the 2026 NSSRN cycle

**DOI:** 10.64898/2026.06.07.26355097

**Authors:** Kellen Squire

## Abstract

**Background:** The emergency department in the United States of America functions as a residual access point for healthcare and social services for populations including rural communities, the uninsured, mental health and addiction patients, and the unhoused. The workforce variable that determines unit function (experience density, the concentration of accumulated clinical judgment within a unit workforce) is not measured in hospital accounting systems.

**Objective:** To document workforce composition changes in U.S. emergency nursing across the 2018 and 2022 cycles of the National Sample Survey of Registered Nurses (NSSRN), and to specify falsifiable predictions for the 2026 cycle.

**Methods:** We analyzed NSSRN public-use files using a current-position ED definition following Castner et al. (2024) and a hospital-bedside-restricted comparator. Variance estimation used jackknife replicate weights for 2018 and Successive Differences Replication for 2022. Burnout was operationalized using the Norful et al. (2023) leaving-reasons proxy across cycles, with sensitivity analysis using the 2022 direct burnout item.

**Results:** A 15-year trajectory (2008–2022) documents progressive experience-density compression: the ED’s 15+ year veteran cohort fell from 41.9% to 28.0% over the decade preceding the pandemic, a contraction of approximately one-third in the senior share and a 19.6% decline in mean experience density, before recovering modestly to 33.3% as veteran nurses remained through the pandemic acute phase, leaving the ED as the youngest hospital setting throughout. Hospital non-ED bedside nurses lost senior tenure between cycles (mean 15.65→14.06 years since first licensure; 15+ year share 43.5%→38.7%), while ED nurses retained their senior tail (mean 11.60→12.58). Burnout endorsement rose sharply in both populations (non-ED 27.3%→46.0%; ED 34.2%→61.2%), with the ED-vs-non-ED gap more than doubling. Controlling for tenure, ED status was not independently associated with burnout in 2018 (OR 1.15, 95% CI 0.83–1.59) but was strongly associated in 2022 (OR 1.92, 95% CI 1.44–2.55; p<.001). The direct burnout item showed a parallel pattern (OR 2.92, 95% CI 1.62–5.28).

**Conclusions:** A pandemic-era setting-specific burnout effect emerged in emergency nursing that workforce-composition controls cannot explain. The 2022 cycle establishes a pre-exit baseline against which the 2026 NSSRN will serve as the falsifiable test of post-Omicron veteran exit. Nursing pipeline replacement lag exceeds the interval before 2026 data arrives; the consequences of inaction fall on populations dependent on ED-based residual access.

## 1. The institution this paper is about

The emergency department functions as the United States’ default twenty-four-hour access point to the healthcare system, and, increasingly, to social services more broadly. As primary care access has declined, mental health infrastructure has eroded, addiction treatment capacity has remained chronically inadequate, rural hospitals have closed, and shelter and protective services have contracted, ED utilization has absorbed the consequences. The empirical signature of this absorption is well documented in rural communities in particular, where median travel distance to access emergency department services rose from 3.3 miles in 2012 to 24.2 miles in 2018 among residents of areas affected by rural hospital closures, with 101 rural hospitals closing between 2013 and February 2020 (U.S. Government Accountability Office, 2020). The same absorption is documented in mental health, where emergency department psychiatric boarding driven by inadequate community and inpatient capacity is now a well-characterized access-block phenomenon (Magarey et al., 2023).

The patient load directed to EDs as residual access point is projected to grow substantially: the Congressional Budget Office estimated that the combined effects of the scheduled expiration of expanded ACA premium tax credits and H.R. 1 coverage provisions would increase the number of uninsured Americans by 16.0 million by 2034, including 7.8 million attributable to Medicaid provisions in H.R. 1 alone (Congressional Budget Office, 2025). Thus, for substantial fractions of the U.S. population (rural communities, uninsured patients, mental health patients, addiction patients, the unhoused, victims of violence) the ED is no longer one healthcare access point among many. It is the **only one available**.

This is not the role the ED was designed to perform. It is the role the ED has *come* to perform because every other system has either contracted or never extended into the populations now relying on it. **ED capacity is therefore not a measure of emergency medical service. It is a measure of residual healthcare and social-service access for the populations the rest of the system has stopped serving.** Degradation of ED workforce capacity therefore manifests not only as longer ED wait times or higher ED-specific adverse-event rates, but as degradation of community access to primary care for the patients who use the ED for primary care, of mental health crisis intervention for the patients with no other crisis service, of pregnancy and reproductive emergency care in states where outpatient options have been restricted, and of the only twenty-four-hour social safety net most U.S. communities now possess.

The residual access function the ED has come to bear was not merely observed; it was *explicitly endorsed* at the highest levels of political discourse as a substitute for universal coverage, a position whose workforce implications were not examined at the time and have not been examined since.

**This paper introduces the construct of *experience density*: the concentration of accumulated clinical judgment available within a unit workforce, proxied here by years since first RN licensure and the tenure distribution of nurses involved in teaching, precepting, or orienting.**

Experience density as deployed here is intended to cover three related forms of clinical judgment: pattern recognition for clinical presentations, awareness of typical failure modes (one’s own past errors and the predictable mistakes of less-experienced colleagues), and the rhetorical capacity to transmit both to novice practitioners. The third element is particularly load-bearing for workforce continuity, as it specifies the mechanism by which experience density reproduces itself across cohorts. A workforce whose senior tail has compressed loses not only the clinical-judgment stock but the transmission infrastructure through which that stock would otherwise be propagated.

Experience density is conceptually distinct from headcount. A unit can maintain stable headcount while losing experience density, if departing senior nurses are replaced by less-tenured nurses. Experience density is also distinct from the composition variable designated as skill mix in the nursing staffing literature. Skill mix refers to the proportion of a workforce composed of registered nurses versus licensed practical nurses, nursing assistants, or other support personnel; the distribution of credential categories within a unit (Griffiths et al., 2026). Experience density, by contrast, operates entirely within the registered nurse category: a unit can maintain both stable headcount and an optimal skill mix while experiencing severe experience density compression, if departing senior registered nurses are replaced by newly licensed registered nurses. Interventions designed to optimize skill mix leave this variable unaddressed and unmeasured.

Hospital accounting systems track and report metrics on headcount continuously and visibly; they do not currently track experience density. The argument developed across this paper is that experience density is the workforce-composition variable most relevant to ED capacity and resilience, and that current institutional metrics systematically obscure changes in it. This institutional blindness has a name in the nursing workforce literature. Hospital staffing models, productivity matrices, and capital-planning frameworks have historically operated under what Aiken and colleagues (2003, 2014) implicitly critiqued: the substitutability assumption that any licensed registered nurse can be exchanged for any other in care delivery models. The educational-composition extension of that critique is now well-documented: nurse education levels translate to patient outcomes in ways the substitutability assumption cannot accommodate (Aiken et al., 2003, 2014; Rochefort et al., 2020).

The present paper extends the critique to a second composition dimension. A 22-year ED nurse and an 18-month ED nurse are both “RN, ED” in the staffing matrix and indistinguishable in the labor line item; the substitutability assumption that makes those matrices function is, however, falsifiable as an empirical claim. The outcome evidence bearing on tenure substitutability comes from the panel literature discussed below; what the present composition data document is the national distribution of that variable in emergency nursing across two NSSRN cycles, and its movement over time.

The staffing literature specific to emergency departments sharpens the point. A 2024 systematic review of nurse staffing and quality of care in emergency departments (Drennan et al., 2024) found that lower nurse staffing levels are associated with longer patient wait times, higher proportions of patients leaving without being seen, increased length of stay, delayed time to medications and therapeutic interventions, and increased risk of in-department cardiac arrest. That review establishes the staffing-level dimension: how many nurses. It also identifies, directly, what that literature has not yet addressed: most of the studies it reviewed did not measure the experience structure of the workforce (length of service, tenure, or specialist emergency qualification); its authors call explicitly for research on the association between workforce experience structure and patient outcomes. Experience density is the structural variable the emergency staffing literature has named as missing and has not yet measured. No prior published analysis has used NSSRN or equivalent national data to characterize longitudinal shifts in ED workforce composition by experience cohort.

The most proximate attempt to operationalize collective RN experience as a distinct staffing dimension alongside education, skill mix, and headcount found the education signal robust but the experience signal null in a single-site setting constrained by low between-shift variance (Rochefort et al., 2020); the authors explicitly call for replication in settings with greater experience variation. The relationship between nursing experience and patient outcomes has a mixed empirical record, and the shape of that record is itself informative. Cross-sectional designs measuring hospital-level mean experience have generally found no association: Aiken et al. (2003) found nurses’ years of experience not a significant predictor of surgical mortality or failure to rescue, and the nine-country RN4CAST analysis likewise found nurse experience not associated with patient mortality (Aiken et al., 2014). Both studies operated on compressed between-hospital variance in mean experience; in Aiken et al. (2003) the between-hospital standard deviation was 2.7 years against a mean of 14.2. High-frequency within-unit panel designs find the opposite. Bartel et al. (2014), using Veterans Administration panel data covering 907,993 patients across 151 acute care units in 76 hospitals, found that increases in average unit tenure significantly reduce residual patient length of stay, with returns that plateau between three and seven years and then rise again through the 10+ year band, while hospital tenure outside the unit was insignificant. Kelly et al. (2026), using ward-day data from an English NHS hospital group, found that one additional year of average hospital-specific experience among degree-qualified nurses reduces the odds of a patient death by approximately 8%, equivalent to adding three-quarters of an additional qualified nurse to the ward, with no corresponding effect for ward- or team-specific experience.

Both found the returns concentrated among registered nurses rather than unlicensed personnel, and Bartel et al. further documented that the departure of an experienced nurse degrades team productivity beyond what the resulting change in average tenure explains, while the departure of an inexperienced nurse does not. Two limitations of that literature bear directly on the present analysis. The panel returns are to unit-specific or hospital-specific experience, which the NSSRN cannot observe, while the proxy used here is general licensure tenure, which is the quantity on which the cross-sectional nulls were estimated. And both panel studies exclude emergency departments from their samples. Experience density is therefore offered here not as a claim that nursing experience has never been linked to outcomes, but as the population-level distribution of a variable measured at the unit and ward level elsewhere, together with the transmission function through which that distribution reproduces itself across cohorts. The outcome consequences of change in that distribution remain a hypothesis, and are treated as such throughout.

This paper documents structural compression in experience density and a pandemic-era setting-specific burnout effect in the U.S. emergency nursing workforce, using the most recent two cycles of the National Sample Survey of Registered Nurses (NSSRN). The hypothesis that the Omicron wave would precipitate a delayed veteran exit from emergency nursing (once acute crisis-retention pressures eased in early 2022) has been a persistent concern among clinicians and workforce analysts, though it does not yet rest on a formal empirical literature. The 2022 NSSRN cycle was fielded with a December 31, 2021 reference date, approximately five weeks before the peak of the U.S. Omicron wave (Iuliano et al., 2022) and before the hypothesized exit window opened. This paper establishes the pre-exit baseline against which the 2026 NSSRN cycle will be the falsifiable test of whether that exit occurred.

The findings are significant regardless of how that test resolves. The pre-exit ED workforce already exhibits compression in experience density and a setting-specific burnout effect that emerged between cycles. The remediation window for these findings closes before the 2026 data is available. The consequences of failing to act, given the residual access function the institution has come to bear, fall not on emergency nursing but on the populations who have nowhere else to go.

## 2. Methods

We analyzed public-use files from the 2018 and 2022 NSSRN, conducted by HRSA and the U.S. Census Bureau. Both cycles use stratified state-level sampling with replicate weights for variance estimation. The 2018 cycle was analyzed using standard jackknife (JK1) variance estimation with 100 replicate weights (RKRNWGTA1–RKRNWGTA100) and scale (R−1)/R.

The 2022 cycle was analyzed using Successive Differences Replication (SDR; Fay & Train, 1995; Ash, 2011), the variance estimation method specified in the 2022 NSSRN Methodology Report (HRSA, 2024). SDR uses 80 replicate weights with a variance coefficient of 4/80; the formula is var(θ₀) = (4/80) Σ(θᵣ − θ₀)². We verified empirically that RKRNWGTA1 in the 2022 public-use file is an alias of the base weight RKRNWGTA (identical values across all hospital-bedside cases) and used RKRNWGTA2–RKRNWGTA80 as the 79 active replicates. In R version 4.6.0 using the survey package (Lumley, 2024), SDR is implemented via the svrepdesign(type="Fay", rho=0.5) specification, which produces mathematically identical variance estimates to the SDR formula above (both yield variance scale = 4/80). Standard errors generated by this specification matched IBM SPSS Complex Samples reference output during preliminary validation following methods described in Kellar and Kelvin (2013). The inferential claims in this paper rest on a small number of pre-specified contrasts (the cross-cycle change in the adjusted ED effect and the within-cycle ED main effects) rather than an exploratory screen across many subgroups; we therefore report uncorrected p-values and do not apply a multiple-comparisons adjustment, which would be inappropriate for a small set of pre-specified hypotheses.

The NSSRN public-use files analyzed here are publicly available from HRSA. Analysis code, including replicate-weight construction, domain estimation (via subset() and svyby() in the R survey package), and the regression specifications reported here, are available in a public repository (https://doi.org/10.5281/zenodo.20576251).

ED nurses were identified using the three current-position criteria of Castner, Zazzera, and Burchill (2024): primary hospital ED or transport setting, non-hospital urgent/emergency/transport setting (2022 only), or Emergency level of care. This specification yields 2,852 ED and 16,063 non-ED cases in the 2022 hospital-bedside comparator. A fourth criterion, Emergency or Trauma Care primary clinical specialty, was examined for definitional robustness and is reported as a sensitivity analysis rather than incorporated into the primary definition: in 2018 it adds no cases, while in 2022 it would add 130 respondents who identify Emergency or Trauma Care as their clinical specialty but whose current position does not otherwise meet ED criteria. Because the setting-specific effect under test is a property of current ED practice rather than of specialty self-identification, the current-position definition is used for all primary contrasts.

All 2018-vs-2022 primary comparisons restrict both ED and non-ED groups to hospital-bedside respondents (those with a valid hospital setting variable), excluding outpatient, education, administration, and research nurses. The broader ED definition (including the 2022-only non-hospital urgent/emergency/transport setting) was used only for count validation against HRSA’s published 2022 NSSRN Dashboard cells, not for primary 2018-vs-2022 contrasts. This isolates ED-specific workforce effects from broader bedside versus non-bedside differences. Cross-cycle non-ED estimates in this paper therefore correspond to hospital bedside nurses outside the ED, not to the broader non-ED nursing workforce.

We verified our sample identification against HRSA’s published 2022 NSSRN Dashboard (HRSA, 2024). Our weighted count of all licensed RNs (4,349,377) matches the dashboard exactly. Our unweighted ED nurse counts by setting variable match HRSA’s published cell counts within rounding tolerance.

PN_BURNOUT in the 2022 PUF is gated on the same-position-throughout-pandemic variable (PN_SAME2020 = 1), restricting the direct burnout question to respondents who held the same primary nursing position from January 1, 2020 through December 31, 2021. Of the 16,467 cases not asked the direct burnout item, 7,753 were not currently employed in nursing and 8,714 were currently employed but had changed primary nursing position during the reference window. These cases appear as system missing in the PUF and were treated as missing in our analyses, after which our weighted endorsement rate (82.3%) matched HRSA’s published figure exactly.

For 2018-vs-2022 cross-year comparison of burnout, we used the same operationalization both years: endorsement of burnout among the leaving-reasons variables, extending the framework developed by Norful et al. (2023). Norful and colleagues operationalized burnout-as-exit endorsement using actual-leaver items only; we extended this to a composite of actual-leaver and considering-leaver endorsement (LE_LVE_BRNOUT and RE_LVE_BRNOUT), to match our research interest in burnout-as-exit risk among the *full* ED workforce population rather than burnout endorsement among the subset who had already exited. The broader composite captures the considering-leaving population that the prediction structure (Section 1) identifies as the cohort at risk for post-Omicron exit. The Norful leaving-reasons operationalization is in active use in the contemporary NSSRN literature; Zhang et al. (2025) applied it across the 2018 and 2022 cycles to compare turnover factors between internationally educated and U.S.-trained nurses, finding burnout a consistent driver of turnover across both groups.

NSSRN does *not* contain a discrete formal-preceptor indicator in the public-use files. We used reported percent time spent "teaching, precepting, or orienting students or new hires" as a proxy for knowledge-transfer workload. Respondents reporting any nonzero time in this category were classified as having teaching/precepting/orienting involvement. This proxy should be interpreted as workload distribution rather than formal preceptor assignment.

## 3. Results

### 3.1 The 15-year trajectory of emergency nursing tenure (2008–2022)

In each of the three NSSRN cycles examined here, U.S. emergency nurses were substantially younger by tenure than other hospital bedside nurses, but the magnitude of the gap and the direction of cross-cycle change differ materially across the trajectory.

In 2008, ED nurses had a mean of 14.43 years since first RN licensure compared to 16.17 years among hospital non-ED nurses, a gap of 1.74 years. The 2008 ED workforce was modestly younger than the rest of hospital nursing but maintained a robust senior tail: 41.9% had 15 or more years since licensure, and 29.7% had 20 or more. The 2008 cycle introduced methodological changes from prior NSSRN administrations (independent state-stratified sampling, regression-based imputation, and differential nonresponse adjustment by age group); the 2008→2018 transition was a more substantial redesign than 2018→2022, and cross-cycle interpretation of 2008 estimates is addressed in Section 9.

For context: the workforce profile above describes the U.S. emergency nursing labor force in the same year that presidential campaigns were deploying the ED as a central argument in the national healthcare debate. The Republican healthcare platform explicitly proposed that ER access constituted effective insurance coverage for the uninsured (Roberson, 2008); the candidates’ approaches to ED crowding and boarding drew standing-room-only attendance at the Society for Academic Emergency Medicine annual meeting and prompted a feature article in Annals of Emergency Medicine (Berger, 2008); Senator Obama introduced the Improving Emergency Medical Care and Response Act of 2007 in direct response to the IOM’s Future of Emergency Care report, a bill that died in committee. The House companion legislation (H.R. 882, 110th Cong., 2007) responded to the IOM report by mandating Medicare reporting of ED boarding times and creating a 10% physician reimbursement supplement for EMTALA-covered services, but contained no provision addressing nursing workforce composition, experience, or staffing quality. Whatever the merits of the policy argument, the 2008 NSSRN data above represents the empirical state of the institution those arguments were predicated upon.

By 2018, however, ED mean tenure had fallen to 11.60 years (95% CI 10.94–12.27), a 2.83-year decline, or a **19.6% loss of mean experience density over a single decade**, while hospital non-ED tenure had moved only modestly to 15.65 years (95% CI 15.39–15.92). The ED-vs-hospital-non-ED tenure gap had widened to 4.05 years, more than doubling. The ED’s 15+ year senior tail had eroded from 41.9% in 2008 to 28.0% in 2018, a loss of 13.9 percentage points: roughly **the 15+ year senior share contracted by approximately one-third in a single decade**. Hospital non-ED nursing, by contrast, retained the bulk of its senior cohort over the same window: the 15+ year share moved only modestly to 43.5%.

In 2022, ED nurses had a mean of 12.58 years (95% CI 11.94–13.22), a modest recovery from 2018, while hospital non-ED tenure had fallen to 14.06 years (95% CI 13.81–14.30). The cross-cycle pattern between 2018 and 2022 inverted: ED tenure rose by 0.98 years, with the 15+ year share recovering from 28.0% to 33.3% (a gain of 5.3 percentage points, consistent with veteran retention during the pandemic acute phase), while hospital non-ED tenure declined by 1.60 years, with the 15+ year share falling from 43.5% to 38.7%.

These share-based comparisons are accompanied by movement in absolute weighted counts that distinguishes compositional change from aggregate workforce contraction. Between 2018 and 2022, hospital non-ED bedside nursing grew from approximately 1.729 million to 1.791 million nurses while its 15+ year population fell from approximately 753,000 to 693,000. Over the same interval, ED nursing moved from approximately 239,000 to 263,000, a change of +24,040 (SE 13,185) that is not individually distinguishable from zero at conventional levels, while its 15+ year population rose from approximately 67,000 to 88,000. Aggregate headcount growth and senior-workforce contraction therefore coexist in the non-ED bedside workforce, and the two settings moved in opposite directions on the senior tail during the pandemic acute phase. This is the empirical content of the distinction between headcount and experience density: a workforce can expand in number while the accumulated clinical judgment available per nurse declines.

The narrowing of the ED-vs-hospital-bedside gap to 1.48 years was not produced by the ED catching up; it was produced by the rest of hospital bedside nursing converging toward the ED’s pre-existing compression. The cross-cycle picture is one of compression on opposite sides of the same gap: the ED held and slightly grew its 15+ year tail while hospital non-ED lost a comparable share of its senior tail (Figures 2 and 4). The within-hospital setting numbers bear out the same pattern.

**Figure 1.**
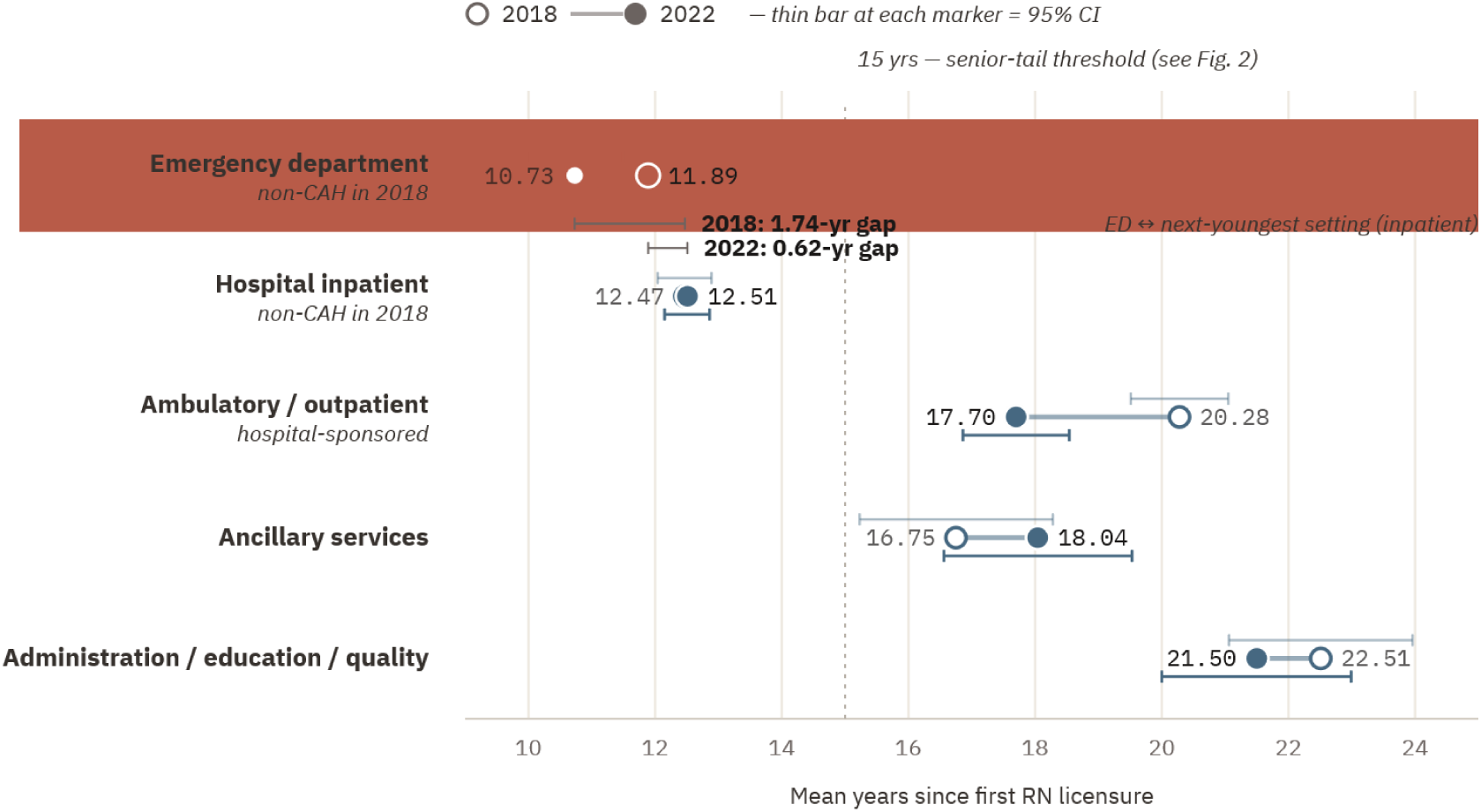
Mean years since first RN licensure by hospital setting, 2018 and 2022. Open markers indicate 2018; filled markers indicate 2022. Thin bars indicate 95% confidence intervals. The emergency department is the youngest hospital setting in both cycles.

**Figure 2.**
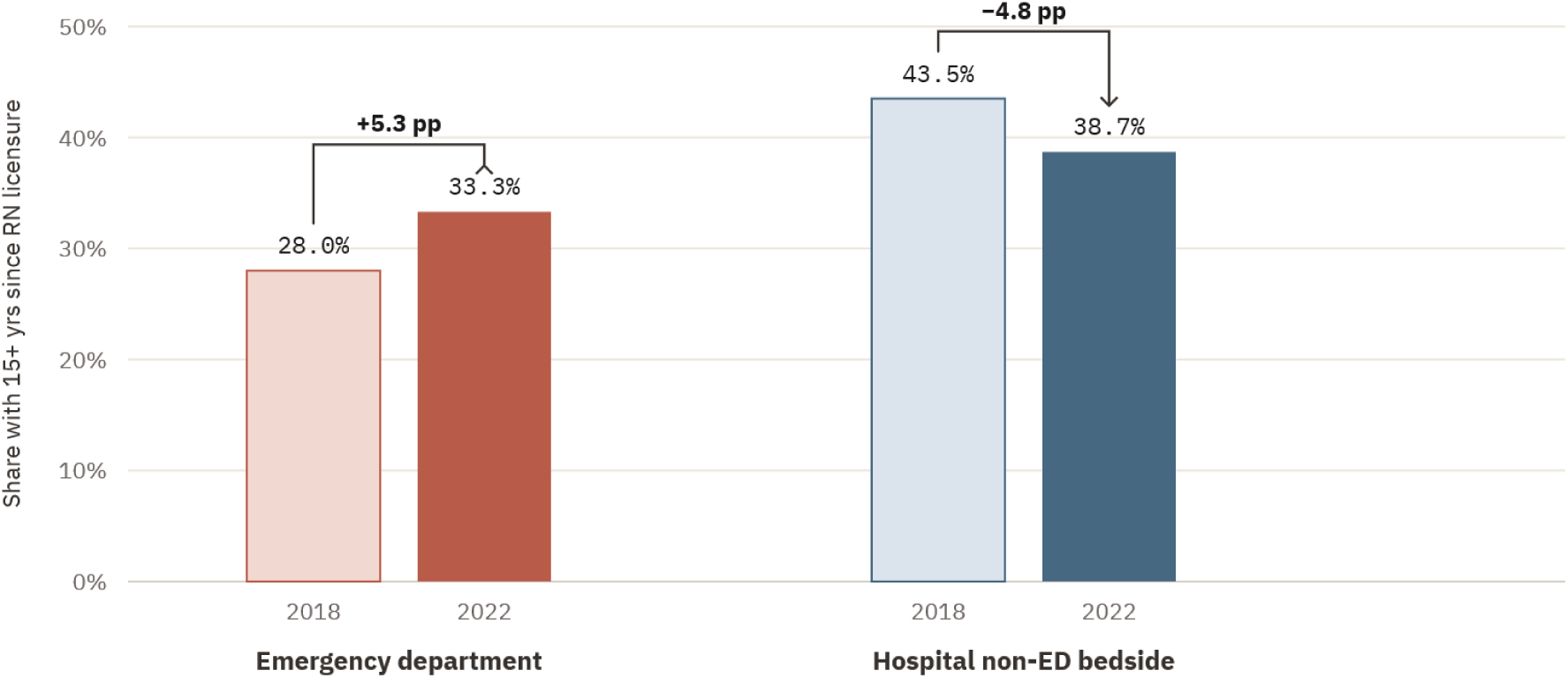
Share of emergency department and hospital non-ED bedside nurses with 15 or more years since first RN licensure, 2018 and 2022. The ED senior tail grew modestly through the pandemic acute phase while the hospital non-ED senior tail declined.

These within-setting means use a finer hospital-setting filter than the primary hospital-bedside comparator and are presented for structural context, not as the primary comparison figures. In 2018, the ED was *the* youngest hospital setting in absolute terms (mean 10.73 years for non-Critical Access Hospital ED, 95% CI 9.98–11.49), followed by hospital inpatient units (12.47 years, 95% CI 12.04–12.89); all other hospital settings (ambulatory clinics, administration, ancillary services, nursing home units, consultative services) exceeded 15 years. The 2022 cycle, which does not separately code Critical Access Hospitals, shows the same pattern: hospital ED at 11.89 years (95% CI 11.10–12.69), hospital inpatient at 12.51 years (95% CI 12.18–12.83), and all other hospital settings between 17.7 and 22.0 years.

**The ED is the youngest hospital setting in both cycles by approximately 1.7 years (2018) and 0.6 years (2022) over the next-youngest setting (Figure 1)**. The structural compression of ED nurse experience is therefore *not* a pandemic artifact; it is a baseline condition the pandemic encountered, and the cross-cycle movement shows the rest of hospital bedside nursing converging toward the ED’s pre-existing compression.

This is the central observation the prediction structure depends on. The 2022 data shows the workforce in a “*holding the line*” phase, before the predicted post-Omicron exit window opens. The ED-side stability (+0.98 years mean, +5.3 percentage points at 15+) is consistent with veteran retention through the pandemic acute phase, though aging-in of the mid-2000s entry cohort across the 15-year threshold is an alternative explanation the present design cannot exclude. The hospital non-ED-side decline (-1.60 years mean, -4.8 percentage points at 15+) is consistent with a leading-indicator hypothesis: the experience-density compression structurally characteristic of emergency nursing may be converging on the broader hospital bedside workforce. The present data document convergence in direction; they do not test transmission between settings.

### 3.2 Burnout endorsement nearly doubled in both populations

Using the Norful leaving-reasons proxy with identical operationalization in both cycles, burnout endorsement rose from 27.3% in 2018 to 46.0% in 2022 among hospital non-ED nurses, and from 34.2% to 61.2% among ED nurses. That a clear majority of the emergency nursing workforce named burnout as a reason for leaving or considering leaving is a catastrophic figure by any operational standard. Both populations’ burnout endorsement rose sharply across the cycle interval, approximately a 68% relative increase in hospital non-ED nurses and 79% relative increase in ED nurses. The cross-cycle change in the ED-vs-hospital-non-ED gap is more striking than the within-population rise: the gap was 6.9 percentage points in 2018 and 15.2 percentage points in 2022. The differential between ED and hospital non-ED burnout endorsement therefore more than doubled across the cycle interval, even as both populations’ absolute rates were rising (Figure 3).

**Figure 3.**
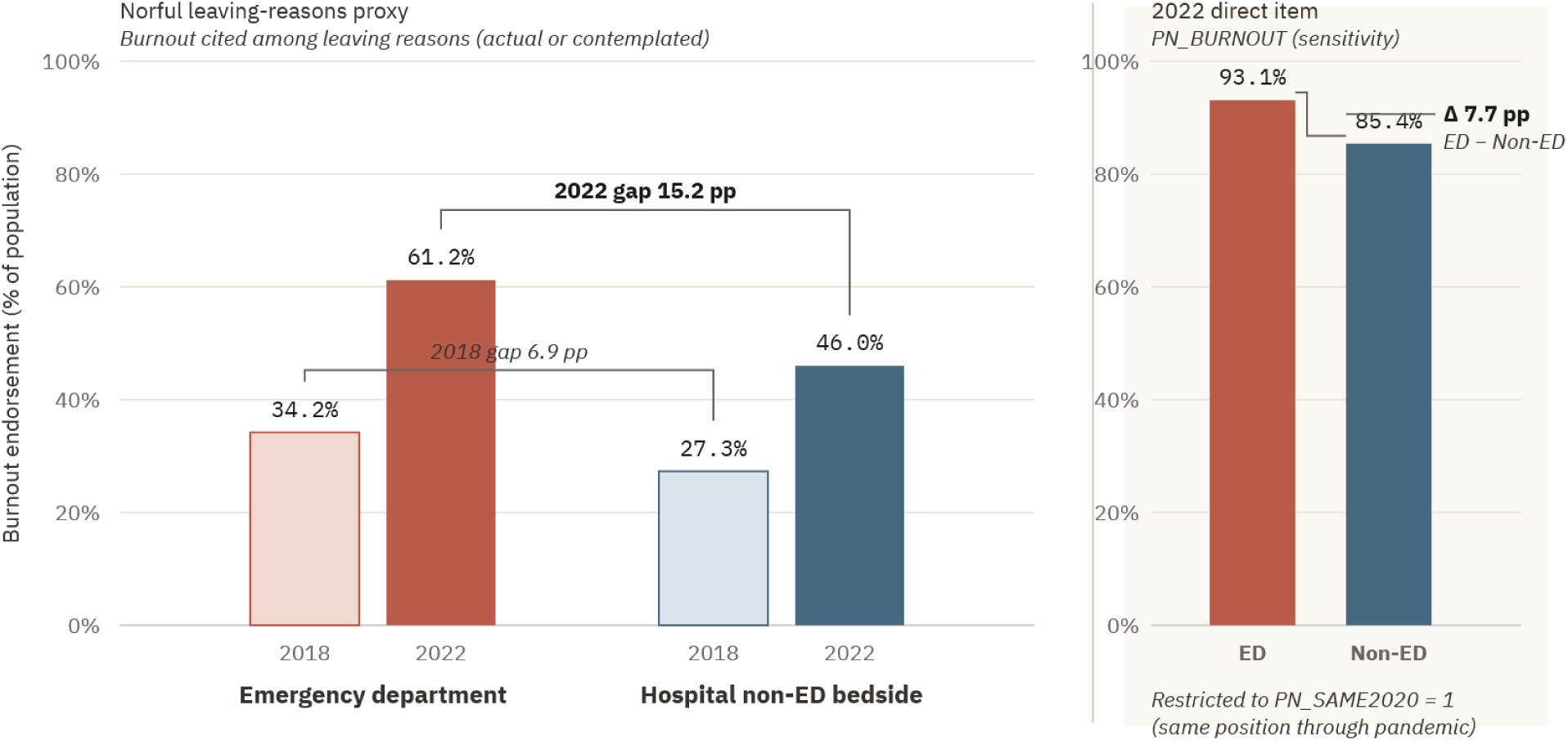
Burnout endorsement by setting and NSSRN cycle. Left panel: Norful leaving-reasons proxy (burnout cited among actual or contemplated reasons for leaving), 2018 and 2022. Right panel: 2022 direct burnout item, restricted to respondents in the same position throughout the pandemic.

**Figure 4.**
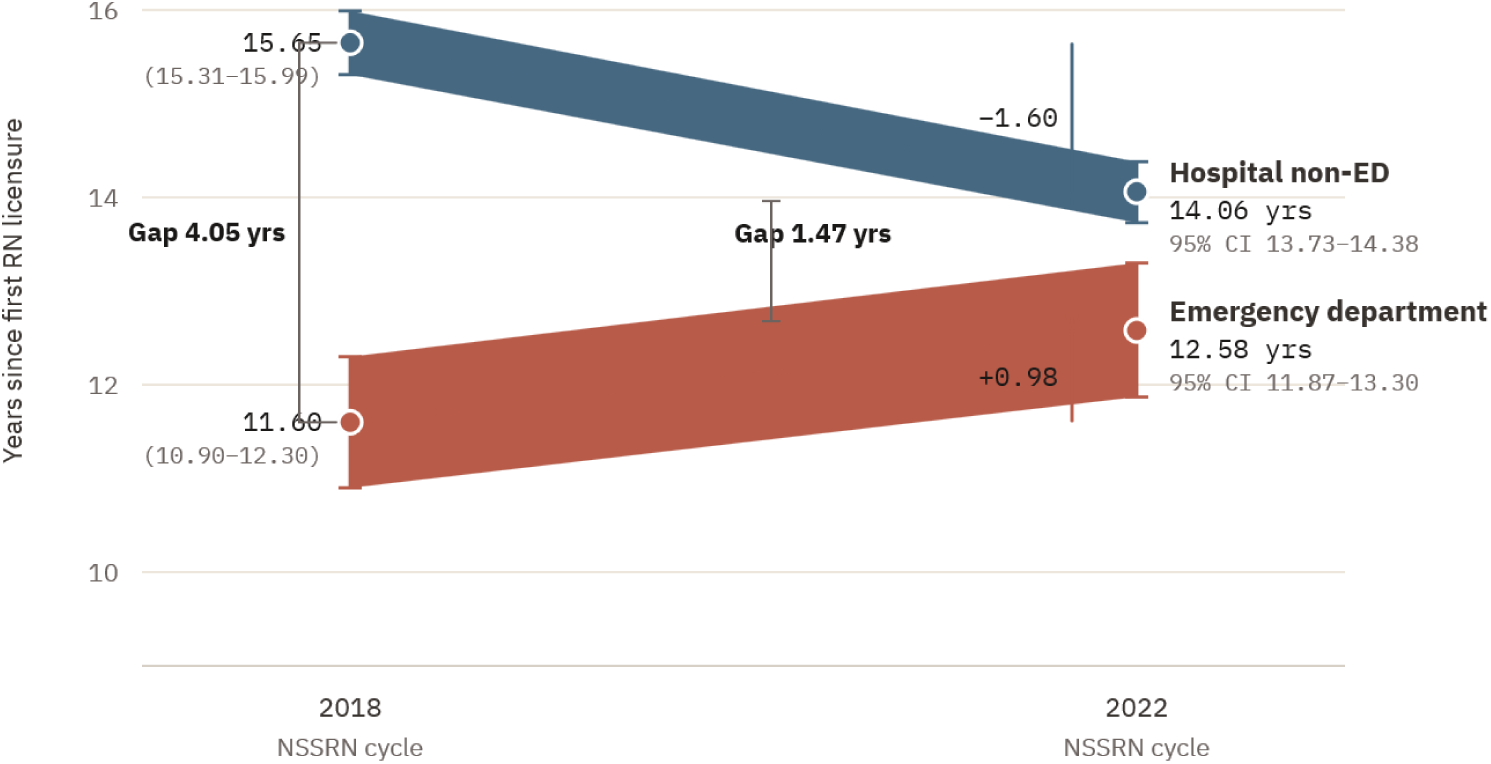
Mean years since first RN licensure, emergency department versus hospital non-ED bedside, 2018 and 2022. Markers indicate weighted means with 95% confidence intervals; shaded bands span the cross-cycle change.

The 2022 direct-question item (corrected for the PN_BURNOUT skip-pattern structure described in Section 2) showed 93.1% of ED nurses and 85.4% of hospital non-ED nurses endorsing any burnout, both near the instrument’s ceiling. The direct-item ED-vs-hospital differential of 7.7 percentage points is narrower than the Norful-proxy differential because the direct item is closer to ceiling in both populations and because the direct item’s denominator (same-position respondents) is a more selected workforce than the Norful proxy’s denominator (the full RN sample).

Read together, the two operationalizations support the same direction of effect. Direct burnout endorsement is approaching ceiling in both populations; the leaving-reasons proxy, which records burnout as a stated cause of either actual or contemplated exit, distinguishes the populations more sharply and shows the ED-vs-hospital gap widening rather than compressing toward ceiling. Whatever produced the 2022 differential is more consequential for stated intent to exit than for endorsement of burnout per se.

### 3.3 The cross-year regression: pandemic-era shift in the ED-burnout relationship

The most substantively important finding concerns how the relationship between ED status and burnout changed between cycles, controlling for tenure.

In 2018, a weighted logistic regression of Norful-proxy burnout on ED status, years since licensure, and their interaction, restricted to the hospital-bedside comparator, found:

- ED status main effect: Wald F = 0.70, **p = .405 (not significant)**
- Years licensed: Wald F = 65.09, p < .001 (OR 0.976 per year, 95% CI 0.971–0.982)
- ED × tenure interaction: Wald F = 1.16, p = .285 (not significant)
- ED odds ratio (controlling for tenure): 1.15, 95% CI 0.83–1.59 (not significant)

In 2022, the identical model on the same hospital-bedside comparator produced:

- ED status main effect: Wald F = 21.17, **p < .001 (significant)**
- Years licensed: Wald F = 208.43, p < .001 (OR 0.961 per year, 95% CI 0.956–0.966)
- ED × tenure interaction: Wald F = 0.41, p = .525 (not significant)
- ED odds ratio (controlling for tenure): 1.92, 95% CI 1.44–2.55

A 2022-only convergent-validity analysis using the direct burnout item (restricted to the same-position subsample to which PN_BURNOUT was administered, and therefore a different estimand than the full-workforce Norful proxy) produced a parallel pattern with an ED main effect Wald F = 13.07 (p < .001), an ED odds ratio of 2.92 (95% CI 1.62–5.28), and a non-significant interaction (Wald F = 1.55, p = .217). Because the ED × tenure interaction was non-significant in both years, the ED odds ratios from the interaction model approximate the adjusted ED effect across the observed tenure distribution; a main-effects-only model yields substantively similar estimates.

A formal cross-cycle test of the change in the adjusted ED-status effect was conducted using a Wald test on the difference of cycle-specific log-odds-ratio estimates. Because the 2018 and 2022 NSSRN cycles draw independent respondent samples, the difference of independent estimates is testable directly; each cycle’s standard error was estimated using its cycle-appropriate variance method (JK1 for 2018, Successive Differences Replication for 2022). This cycle-specific approach, rather than a pooled cross-cycle model, follows the precedent of Samson et al. (2025), who analyzed the 2018 and 2022 NSSRN cycles separately owing to sample-frame changes between cycles. The 2022 log odds ratio exceeded the 2018 log odds ratio by 0.5134 (SE 0.2178), z = 2.358, two-tailed p = .018. Equivalently, the adjusted ED-vs-non-ED odds ratio for burnout-as-exit endorsement was 1.671 times higher in 2022 than in 2018 (95% CI 1.090 to 2.560). The cross-cycle change in the setting-specific effect therefore exceeds what would be expected from sampling variability alone.

A traveler-adjusted sensitivity analysis tested whether the 2022 ED main effect on burnout-as-exit endorsement was driven by short-position-tenure composition, operationalized via traveler status (PN_TRAVEL). Travelers accounted for 7.4% of non-ED and 7.3% of ED nurses in the hospital-bedside comparator, with substantially younger tenure profiles (mean licensure tenure 8.1 and 9.5 years among non-ED and ED travelers respectively, versus 14.5 and 12.8 years among permanent staff). After adjustment for traveler status, the ED main effect on Norful-proxy burnout remained highly significant (OR 1.79, 95% CI 1.49 to 2.15, p < .001), a modest attenuation from the unadjusted 1.92 consistent with traveler status carrying part but not the bulk of the variance the ED main effect accounts for. The traveler-status main effect was itself significant and ran in the opposite direction: travelers endorsed burnout-as-exit at lower rates than permanent staff (OR 0.68, 95% CI 0.50 to 0.91, p = .010), a finding inconsistent with the popular framing that travel nursing produces elevated burnout and more consistent with structural buffering: nurses who hold the option to leave at contract end may be less likely to endorse leaving-reason proxies that capture cumulative exit motivation.

The cross-year contrast is the finding. In 2018, the ED’s elevated crude burnout endorsement was largely explained by the ED workforce’s lower licensure tenure profile; after adjustment for tenure, ED status was not independently associated with burnout. By 2022, ED status was independently and substantially associated with burnout endorsement after adjustment for tenure. The interaction term was non-significant in both years, indicating that the protective effect of tenure operated similarly in both settings. What changed between cycles was the setting-specific effect of working in the ED.

Stated carefully: the pandemic-era ED appears to have become independently associated with burnout endorsement after adjustment for tenure, in a way the pre-pandemic ED was not. The 2026 NSSRN cycle reframes the direct burnout item away from pandemic-specific attribution (Section 5.1); the 2018–2022 contrast documented here is likely the only NSSRN cross-cycle observation in which the emergence of this setting-specific effect can be measured against the pandemic-framed instrumentation that captured it. We address mechanism hypotheses in the Discussion (Section 4).

### 3.4 The EMS-to-RN pipeline

In 2022 (the first NSSRN cycle to include a discrete prior-EMT/paramedic-employment variable) 15.4% of ED nurses (95% CI 12.8–18.5%) reported having been employed as an EMT or paramedic before completing their first RN degree, compared to 3.5% of non-ED nurses. Emergency nurses were 4.4 times more likely than non-ED nurses to have entered nursing through the EMS pipeline. The 2018 cycle does not separately code EMT/paramedic prior employment, preventing direct cross-year testing of pipeline composition.

### 3.5 Knowledge-transfer involvement and the leading-indicator hypothesis

The leading-indicator hypothesis advanced in Section 3.1 (hospital non-ED bedside nursing converging toward ED-baseline tenure compression) extends into the knowledge-transfer subpopulation. Among nurses reporting any teaching, precepting, or orienting time, ED nurses were substantially less experienced by RN licensure tenure than non-ED nurses in both survey cycles. In 2018, ED nurses with knowledge-transfer involvement averaged 11.04 years since first RN licensure (95% CI 10.20–11.88), compared with 15.73 years (95% CI 15.39–16.08) among non-ED nurses (gap 4.69 years, p < .001). In 2022, the corresponding means were 11.58 and 14.36 years (gap 2.78 years, p < .001).

The narrowing of the gap between cycles is driven primarily by decline on the non-ED side rather than improvement on the ED side. ED knowledge-transfer tenure was approximately stable (11.04 to 11.58 years). Non-ED knowledge-transfer tenure declined by 1.37 years (15.73 to 14.36). The proportion of non-ED knowledge-transfer nurses in the highest tenure band (25+ years since licensure) fell from 24.9% in 2018 to 13.4% in 2022, while the corresponding ED proportion fell from 11.3% to 6.9%. The structural pattern observed in emergency nursing (knowledge transfer carried disproportionately by an early- and mid-career workforce) is increasingly visible across non-ED nursing as well, on a delayed but parallel trajectory.

Nurses who remain in emergency nursing beyond a decade are not simply more experienced versions of their early-career colleagues. They are a psychologically selected cohort whose profile differs in measurable ways from both early-career peers and from those with moderate tenure who intend to leave (Winters, 2019). This selection means that compositional loss at the senior tail is not recoverable by recruitment alone; the replacement pool does not contain the same workforce.

The experience density collapse documented here has an interprofessional dimension that extends beyond the nursing workforce. Experienced nurses have historically served as informal clinical educators for new residents and interns, a knowledge transfer function documented in ICU and obstetric settings (Petri et al., 2024; Doja et al., 2021) and now the subject of formal intervention programs in emergency medicine precisely because the organic version can no longer be assumed (Doodlesack et al., 2024). That a structured curriculum is required to restore what once occurred as a matter of course is itself evidence of baseline deterioration. The simultaneous loss of experienced nurses from the workforce and the erosion of the interprofessional channel through which their clinical judgment once propagated to new physicians compounds the safety failure beyond what experience density numbers alone measure.

**Experience density in knowledge-transfer roles declined in hospital non-ED nursing between cycles and remained approximately flat in the ED (11.04 to 11.58 years)**, with the ED operating below the rest of the hospital throughout and the gap between them narrowing **(Figure 5).**

**Figure 5.**
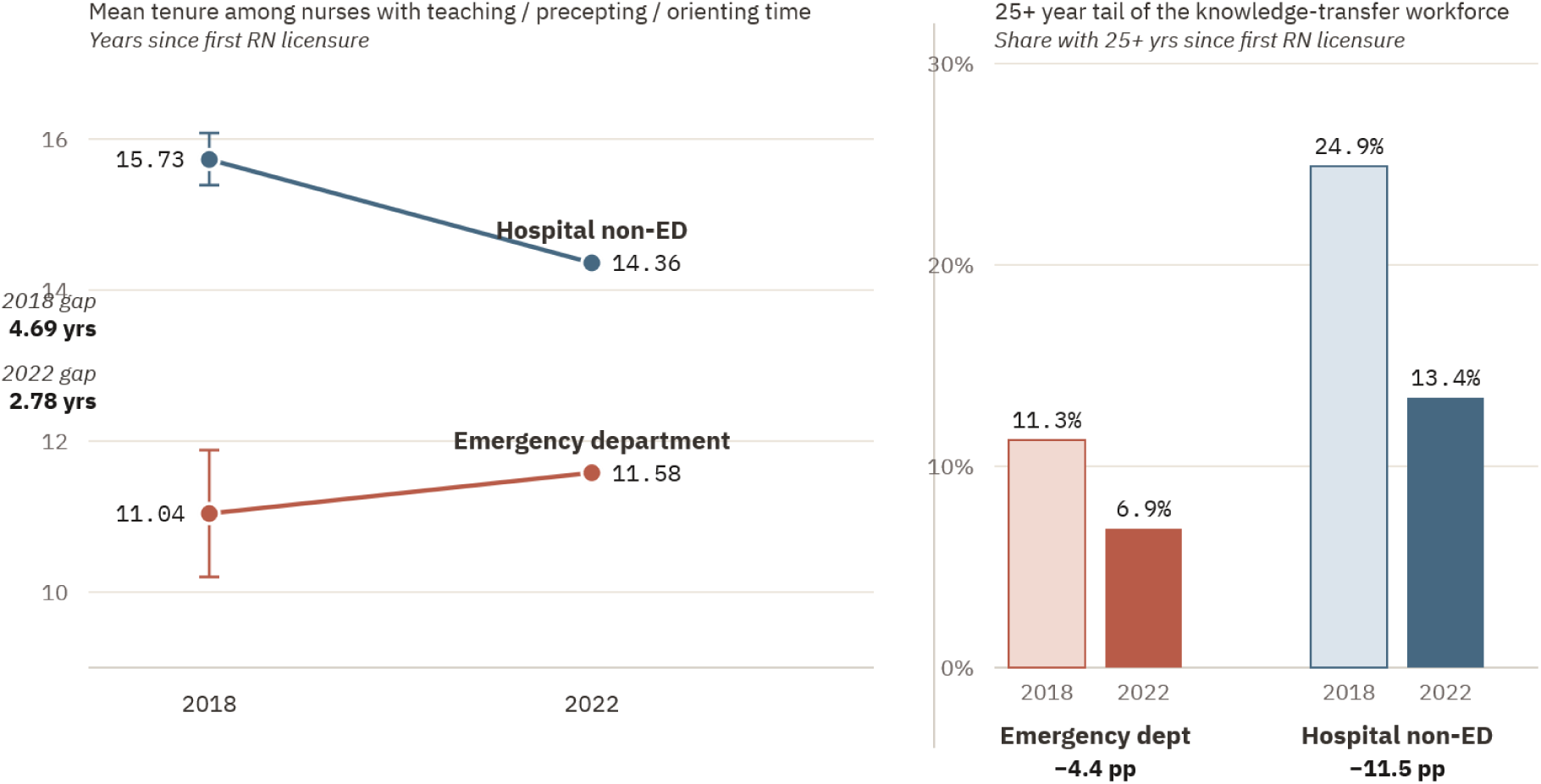
Tenure among nurses reporting teaching, precepting, or orienting involvement, 2018 and 2022. Left panel: mean years since first RN licensure with 95% confidence intervals. Right panel: share of the knowledge-transfer workforce with 25 or more years since first RN licensure.

### 3.6 What the data rules out

Mean self-reported hours worked per week were essentially identical between ED and non-ED nurses in both cycles (2018: 37.49 vs 37.52; 2022: 35.38 vs 36.12). The elevated burnout differential observed in 2022 was not explained by mean weekly hours quantity. Whatever produced the setting-specific burnout effect operated within the same hours envelope as non-ED nursing; the mechanism is *not* raw work volume.

This rules out the simplest “ED nurses just work more” framing, but does not address shift-distribution patterns, mandatory overtime clustering, missed-break frequency, or workload intensity within work hours, all of which remain plausible mechanisms.

The 2022 NSSRN also documents a workforce-instability indicator the regression analyses do not capture: among 41,361 RNs currently employed in nursing on December 31, 2021, 8,714 (21.1%) reported having changed their primary nursing position between January 1, 2020 and December 31, 2021. This is the population the direct burnout question excludes by design (Section 2). It is also a population for whom the Norful leaving-reasons proxy may underestimate burnout if position change was itself a partial exit response that did not reach licensure-status exit. The aggregate position-change rate during the pandemic acute phase is therefore an additional empirical signal of workforce instability whose attribution to pandemic-era stress is no longer testable in the 2026 cycle (Section 5.1).

## 4. Discussion

The empirical pattern documented in Section 3 supports three substantive conclusions about the U.S. emergency nursing workforce.

**First, the ED is operating with structurally lower experience density than the rest of hospital nursing across all cycles examined, and this compression had already deepened over the decade preceding 2018 rather than appearing as a 2018 baseline condition or emerging during the pandemic period** (Section 3.1). The contrast is sharpest at the senior-preceptor tier: at the 25+ years of licensure threshold, used here as a marker of deeply accumulated clinical experience rather than as a formal preceptor designation, the ED workforce carried 12.5% and 13.8% across the 2018 and 2022 cycles, against a non-ED hospital workforce that fell from 25.1% to 19.4% over the same period. **The ED has been operating without a robust senior-preceptor cohort throughout the study period; hospital non-ED bedside has now lost nearly a quarter of its senior cohort over four years and is converging toward the ED’s structurally compressed baseline.** The within-hospital setting structure documented in Section 3.1, with the ED as youngest hospital setting in all cycles, hospital inpatient closely behind, and all other hospital settings (ambulatory, administration, ancillary, consultative) substantially older, locates this compression *specifically* at the hospital bedside, distinct from the broader non-bedside nursing workforce that retains its senior cohort.

**Second, the ED-burnout relationship changed materially between cycles in a way that workforce-composition controls cannot account for.** The 2018 model is consistent with an interpretation in which ED-elevated burnout was largely a tenure-composition phenomenon: junior nurses are more burned out everywhere, the ED has more junior nurses, the ED appears more burned out. By 2022, ED status was independently associated with burnout endorsement after adjustment for tenure, with odds ratios in the 1.9–2.9 range depending on the burnout measure used. The interaction term was non-significant in both years, indicating that tenure protected nurses similarly in both settings; what changed was a setting-specific effect of working in the ED.

The 2018→2022 recovery in ED senior tenure (Section 3.1) occurred during a period when ED nurses’ self-reported reasons for remaining in their positions, as documented in a 2025 scoping review of 93 retention and turnover sources, prominently included a novel and dominant theme of moral obligation to remain despite personal risk (Boulton et al., 2025). The empirical signature documented here is consistent with exit-suppression under perceived moral constraint: senior nurses who, under non-pandemic baseline conditions, might have transferred out of ED practice or retired, instead remained through the acute pandemic phase. The cost of that retention, on this reading, registered not as immediate exit but as the elevated burnout-as-exit endorsement the same cohort exhibits in the 2022 data: the leading edge of a delayed rather than averted exit signal. The cross-sectional NSSRN proxy cannot independently establish causation, but the cross-cycle pattern is consistent with this interpretation, and the 2026 cycle provides a falsifiable test: if exit-suppression under moral constraint is the mechanism, the post-acute-phase release of that constraint should now be visible as accelerated senior exit.

**Third, the empirical findings are silent on mechanism but their workforce-composition prerequisites support several testable hypotheses.** One we wish to articulate explicitly, because it follows directly from the experience-density framing: a less-experienced nursing workforce facing higher per-patient acuity than their training prepared them for may default to wider workups, lower admission thresholds for ambiguous presentations, and longer per-patient ED dwell times, defensive utilization patterns that consume bed-hours, drive boarding, compound crowding, and amplify the very environmental conditions that produced the burnout response in the first place.

The behavioral premise underlying this hypothesis is not speculative. Levis-Elmelech et al. (2022), analyzing more than eighteen thousand triage records at a single tertiary center, found that the direction of triage error is experience-graded: less-experienced nurses were measurably better at flagging genuinely high-acuity patients but systematically *over*-triaged ambiguous ones, while experienced nurses were measurably better at identifying which patients were *not* sick, the authors attributing the novice pattern to triage anxiety and lower decision confidence. This asymmetry is the operationally consequential one. Over-triage of the ambiguous presentation is the resource-expensive error: it is the decision that orders the wider workup and claims the bed. The capacity to confidently classify a patient as low-acuity, and conserve those resources, is precisely the capacity that Levis-Elmelech locates in experienced nurses and that an experience-density collapse strips from the workforce. Their data establish that this classification turns measurably on the experience of the nurse making it, but the study stops at the classification itself; it does not follow that decision downstream into workup intensity, admission, dwell time, or boarding. Levis-Elmelech thus document the first link in the chain (and the direction of its loading) without measuring the magnitude of what follows; the downstream utilization cascade remains untested here. NSSRN does not measure ED throughput, workup intensity, or admission decisions. The workforce-composition prerequisites for this hypothesis are documented here; its empirical test requires linkage to operational data this paper does not access. We note that the hypothesis, if confirmed, would close a feedback loop in the existing ED boarding and crowding literature, which has generally treated staffing as exogenous to throughput conditions: under the experience-density framing, low experience density would produce defensive utilization, which drives boarding and crowding, which amplify burnout and accelerate experience exit, which lowers experience density at the next cycle.

The downstream consequences of this feedback mechanism are now quantifiable at the national level in international surveillance data: in Canada, waiting for an inpatient bed accounted for an average of 59% of an admitted patient’s total emergency department stay in 2024–2025, a 45% increase in median bed-wait time since 2018–2019, with Canada recording the highest acute care bed occupancy among 32 OECD countries at 91%, against an OECD average of 71% (Canadian Institute for Health Information, 2026b). Emergency departments report the highest nursing overtime rates among hospital units in Canadian national data at 11% in 2023–2024, more pronounced in rural hospitals than urban, the operational signature of the same workforce strain the NSSRN burnout data documents in the U.S. context (Canadian Institute for Health Information, 2026b). These indicators do not test the feedback mechanism described above, but they establish that the same pattern of strain is present in a universal-coverage system, where the insurance variable often invoked to explain U.S. ED pressure is absent.

An analogous workforce-capacity dynamic has been documented in radiology, an adjacent acute-care specialty in which strain is directly observable in throughput metrics: imaging interpretation turnaround times for Medicare outpatients rose 113 percent between 2014 and 2023, with computed tomography turnaround increasing 318 percent, and the abrupt 2022– 2023 acceleration was interpreted by the authors as evidence that the radiology workforce had reached maximum capacity (Christensen et al., 2026). Because ED throughput is partly gated by imaging turnaround, this finding both corroborates the feedback-loop structure proposed here and identifies one concrete pathway through which adjacent-specialty workforce strain lengthens ED dwell times independent of nursing staffing. Other plausible mechanisms, such as boarding intensification, crowding, workplace violence, moral distress, prolonged critical-care holds, COVID exposure cumulative effects, staffing instability, all remain candidates and are not mutually exclusive.

Furthermore, the cross-year contrast does not establish causation. We cannot conclude from these data that the pandemic caused the ED-burnout shift documented here, only that the relationship between ED status and burnout changed during a period in which the pandemic occurred. Establishing causation requires additional data sources and study designs beyond this paper.

No state health agency currently tracks experience density as a workforce metric separate from headcount in its licensed nurse population. The gap is not incidental: a shortage defined by number-of-nurses generates different policy responses than one defined by experience distribution, and the former framing will systematically understate the problem the latter reveals. Prospective tracking of experience density (reported next to headcount rather than in place of it) is technically feasible using existing licensure data infrastructure and is the measurement prerequisite for any policy intervention designed around workforce composition rather than workforce volume.

These workforce-composition findings are corroborated by independent industry and academic data. Auerbach et al. (2024), analyzing Current Population Survey data through 2023, document that between 2018–19 and 2022–23, hospital employment among RNs aged 40 and older declined by 4.9 percent, while hospital employment among RNs younger than 40 grew by 9.2 percent, a compositional shift that (like the NSSRN-documented experience-density compression) is invisible in aggregate RN headcount projections.

Industry survey data from NSI Nursing Solutions, Inc. (2026), based on 527 hospitals and 262,405 RNs, places annual ED nurse turnover at 20.7 percent (well above the national RN average of 17.6 percent) and reports that the five-year cumulative turnover rate for ED RNs is 113.6 percent, a volume of separations equivalent to **more than the entire ED RN headcount over that period**. Independent corroboration through Current Population Survey methodology and through industry retention survey methodology converges on the workforce dynamic the NSSRN data documents. The pressure is visible at both ends of the experience distribution: Church et al. (2025), analyzing the same 2018 and 2022 NSSRN cycles, found burnout-driven turnover intent elevated among newly-licensed registered nurses, indicating that the junior cohort meant to replenish the senior tail is itself at heightened burnout-attributable exit risk — a compromise of the replenishment mechanism on top of the senior-tail compression documented here.

## 5. The 2026 NSSRN cycle as the predictive test

The structural baseline established here predicts a specific empirical pattern in the 2026 NSSRN cycle, conditional on the post-Omicron veteran exit hypothesis being correct.

If the prediction holds, the 2026 cycle should show reduction in the proportion of ED nurses with 15+ years since first licensure compared to 2022, decline in absolute weighted ED nurse count if replacement hiring did not match exit volume, further narrowing of the senior tail in non-ED knowledge-transfer cohorts as the structural pattern propagates, and a burnout endorsement pattern shaped by the offsetting forces of selection (those who stayed are by definition the most resilient) and compression (a smaller workforce is more loaded per nurse). If the prediction does not hold, the 2026 cycle should show roughly stable ED workforce composition relative to 2022.

The prediction is therefore falsifiable, with the 2026 NSSRN public-use file as the test instrument.

The 2026 cycle measures more than nursing workforce composition. It measures whether the institution that has absorbed residual access functions for an increasing share of the U.S. population is structurally capable of continuing to do so.

### 5.1 Risk to the test instrument

The predictive value of this baseline depends on continuity in federal workforce surveillance. The 2026 NSSRN cycle, currently in field with a January 2026 first mailing and a planned June 2026 closeout (U.S. Census Bureau, OMB Control No. 0607-1002), introduces three changes that bear on the falsifiable test described above.

#### Temporal positioning of the 2022 reference date

The 2022 NSSRN reference date of December 31, 2021 precedes the peak of the U.S. Omicron wave (January–February 2022) by approximately five weeks. The Omicron wave produced the highest weekly U.S. SARS-CoV-2 case counts and among the highest weekly death counts of the entire pandemic (Iuliano et al., 2022), representing the largest concentrated occupational exposure event for the U.S. healthcare workforce. The workforce composition documented in this paper therefore represents a measurement taken immediately before, rather than during or after, the wave hypothesized to drive post-acute-phase exit. The 2026 cycle is the first NSSRN cycle to measure the workforce state after the predicted post-Omicron exit window.

#### Removal of pandemic-exit instrumentation

The 2026 cycle removes Section J of the 2022 questionnaire ("Nursing During the Coronavirus Pandemic") in its entirety (Appendix A, OMB Supporting Statement A, 2025). The retained burnout instrumentation expands relative to 2022 (the 2026 cycle adds a four-item burnout severity scale, a burnout-frequency item, and a burnout-cause attribution battery in addition to the direct burnout endorsement item) but the items specifically asking whether respondents left nursing between March 2020 and December 2021, the reasons for leaving (including pandemic-specific causes), and whether they intended to return are eliminated. The Norful leaving-reasons proxy used in our cross-cycle analysis is not in the documented changelog and appears retained; whether its skip-pattern denominators remain comparable across cycles will require verification against the released 2026 PUF.

Active-license sampling means nurses who left nursing during the pandemic but retained licensure remain in the 2026 sampling frame; they are not, however, identifiable as pandemic-era exits in the absence of Section J’s retrospective items. The cohort of nurses who left in response to Omicron-acute-phase exposure and did not return is therefore observable in aggregate (through population-level headcount, age, and tenure-distribution shifts measurable in 2026) but not as an identifiable subpopulation with attributable causes for exit. The longitudinal-causal observation specifically foreclosed by this combination of survey timing and instrumentation change is the attribution of post-2022 workforce attrition to the pandemic event that may have produced it.

#### Executive modification of federal statistical surveillance

The 2026 NSSRN cycle is conducted under Executive Order 14168 (2025), explicitly cited in the cycle’s OMB Supporting Statement as governing federal sex and gender data collection. This constrains the demographic instrumentation available in 2026 and subsequent cycles relative to 2018 and 2022. The constraint does not affect the validity of the analyses presented here, but (independent of the substantive merits of any specific directive) it establishes a precedent for executive modification of the measurement framework underlying federal workforce surveillance. That precedent is not isolated: in June 2026 the Department of Commerce issued Department Administrative Order 216-26 (U.S. Department of Commerce, 2026), prohibiting noise infusion as a disclosure-avoidance method across all Census Bureau and Bureau of Economic Analysis statistical products and establishing coarsening and, as a last resort, suppression as the permitted alternatives. Within approximately sixteen months, then, two executive administrative instruments—spanning two federal statistical agencies and two distinct methodological domains, demographic categorization and statistical disclosure limitation—modified the methodological substrate of federal statistics. Future longitudinal interpretation of NSSRN data must account both for the resulting discontinuity in measurement framework and for the demonstrated, and now recurring, possibility of further such modifications across cycles. Where disclosure-avoidance options narrow to coarsening and suppression, moreover, the variables most vulnerable to loss are precisely those resting on thin cells, including the small high-tenure subpopulations on which the experience-density measures developed here depend.

#### What the 2026 cycle can and cannot test

The 2026 NSSRN will permit testing of the workforce-composition predictions specified above: whether the proportion of ED nurses with 15+ years since first licensure declines from the 2022 baseline; whether absolute weighted ED nurse count declines if replacement hiring did not match exit volume; whether the senior tail in non-ED knowledge-transfer cohorts narrows further; and whether the EMS-to-RN pipeline composition documented for 2022 changes. The Norful leaving-reasons proxy, conditional on instrumentation continuity, will permit cross-cycle comparison of burnout endorsement. The 2026 cycle will not permit attribution of observed workforce changes to pandemic-era exposure at the individual level, nor will it permit identification of the cohort of nurses whose exit was driven specifically by pandemic occupational experience. Workforce composition trajectories remain testable; the causal chain connecting them to the event hypothesized to have driven them is severed at the 2022 cycle.

Continued public availability of the 2026 cycle in a methodologically continuous form is itself a matter of healthcare system surveillance integrity. Any further methodological discontinuity, delay in release, or suppression of comparable workforce variables would compound the limitations described here. Given the policy stakes documented in Section 1, the surveillance infrastructure that permits empirical evaluation of post-pandemic workforce-composition changes is not separable from the workforce changes themselves; both are operating under the same set of structural constraints.

## 6. The training pipeline lag and self-reinforcing capacity degradation

Nursing workforce remediation operates on a horizon of roughly four to seven years, treated here as a planning estimate rather than an empirically established interval: New BSN matriculation to licensure: four years. New-grad-to-safe-to-precept: another two-to-three. New-grad-to-able-to-anchor-an-emergency-department-overnight: another two-to-three on top of that.

The implication: if the 2026 NSSRN confirms substantial post-Omicron veteran exit, **the remediation window for that exit closed before the data confirming the exit was published.** A response initiated in 2026 would not produce newly licensed nurses until roughly 2030, preceptor-capable replacements until approximately 2032–2033, or experienced ED anchors until the mid-2030s. Restoration of pre-pandemic experience density across the hospital nursing workforce as a whole would not occur until 2035–2040 at the earliest, which is still an ambitious timeline.

What happens during the lag is no longer hypothetical. The U.S. literature on ED boarding and crowding documents a self-reinforcing pattern in which longer waits worsen patient acuity, sicker patients require longer treatments, longer treatments consume capacity, reduced capacity produces longer waits, and each cycle yields a system further from baseline than the one before (Pines et al., 2009; Singer et al., 2011; Sun et al., 2013). This dynamic operates at the level of the unit and the shift, not the institution and the year, which is why it is invisible to quarterly headcount metrics but visible to the bedside.

The pattern is consistent with what clinician-preference data has established: in a 2021 multi-site survey, nurse burnout was significantly associated with both nurse and physician turnover at the hospital level, with turnover rates 5–8 percentage points higher in hospitals at the 75th percentile of nurse burnout, dissatisfaction, and intent-to-leave compared to the 25th percentile (Aiken et al., 2023). The during-lag dynamic does not require speculation; the empirical relationship between burnout, turnover, and workforce stability is documented at the hospital level in peer-reviewed work.

The argument for acting on the pre-exit baseline is therefore an asymmetric expected-value argument:

- **Act now, prediction wrong:** society over-invests in nurse retention. Given that current retention investment is demonstrably below replacement-cost levels (Section 7), even substantial over-investment is unlikely to exceed the operational savings of reduced turnover.
- **Don’t act now, prediction wrong:** no consequences. Workforce stable.
- **Act now, prediction correct:** self-reinforcing degradation is blunted earlier, residual access function preserved, fewer adverse events for the populations dependent on it.
- **Don’t act now, prediction correct:** progressive capacity degradation through the late 2020s falling on the institution that has become the residual healthcare and social-service access point. Replacement cohorts arrive in the early 2030s. The intervening years’ costs fall on rural communities, uninsured patients, mental health patients, addiction patients, the unhoused, and victims of violence.

The expected-value calculation supports acting on the pre-exit baseline rather than waiting for post-exit confirmation, regardless of one’s prior probability on the original prediction. **Anyone arguing against the pre-exit response must explicitly accept the downside**. They must argue that the populations who currently rely on the ED for residual access do not warrant the cost of action, or must deny the residual access pattern itself, which is empirically documented.

## 7. Why current institutional structures are not acting

Hospital systems have not responded to documented workforce compression with retention investment commensurate to documented replacement cost. Published estimates place the cost of replacing a tenured RN at $50,000–$80,000 per nurse, including recruitment, three-to-six months of orientation at full salary with limited independent productivity, preceptor time pulled from existing staff, ramp-up-period error rates, and turnover-driven floor instability that triggers additional departures (NSI Nursing Solutions, Inc., 2026; Bae, 2022). For a 5-year-tenure ED nurse, **the replacement cost meaningfully exceeds the cumulative cost of pension contributions, retention bonuses, or wage adjustments that would have prevented the departure.**

The hospital labor literature documents the same structural pattern. Hospital consolidation has been associated with slowed wage growth for hospital workers with industry-specific skills, with the effect particularly pronounced in markets with weaker union presence (Prager and Schmitt, 2021). Greater hospital-system concentration is associated with lower real hourly RN wage growth in smaller metropolitan areas (Allegretto and Graham-Squire, 2023). National wage trend data show that **RN wages experienced the smallest annual inflation-adjusted growth of any healthcare occupation** studied between 2012 and 2023 (0.51 percent), while nursing assistants experienced the largest (1.48 percent; Ratliff et al., 2025).

Hospital financial resources, including pandemic-era subsidies, have not been associated with higher nurse staffing levels; in the pandemic period specifically, **hospitals with greater financial resources actually maintained lower nurse hours per patient day than hospitals with fewer resources** (Chong et al., 2025). This forecloses the explanation that low nurse staffing is mainly a function of financial constraint and points instead to allocation decisions internal to hospital management. International evidence from elective surgery waiting times across OECD countries similarly suggests that post-pandemic capacity degradation did not systematically relate to pre-pandemic health spending, physician density, or acute bed capacity (Siciliani et al., 2026), reinforcing the structural rather than financial nature of the problem this paper documents.

Meanwhile, nonprofit hospital executive compensation has continued to rise. CEO pay at U.S. nonprofit hospitals is associated more closely with hospital size and financial performance than with quality outcomes or community benefit (Jenkins et al., 2024; Jenkins et al., 2025), and recent reporting has documented average executive compensation increases of approximately 41% at one major integrated delivery network during a period of record system revenue (North Carolina Health News, 2025). Multiple major integrated delivery networks have, between 2022 and 2026, frozen or eliminated nursing pension benefits, suspended retirement contributions, proposed wage freezes or below-inflation raises, or reduced benefit structures, even as system revenues, executive compensation, and capital expansion have continued to grow. Public examples include pension freezes at Intermountain Health and BJC Health System (Becker’s Hospital Review, 2025), retirement-contribution suspensions at Marshfield Clinic (Marshfield Clinic Health System, 2023), and benefit and wage disputes at multiple academic medical centers and community hospitals during contract negotiations.

**Money spent on headcount does not fix this. The structural problem is experience density.** Hiring more first-year nurses to replace exiting fifteen-year nurses keeps the headcount line item flat while the experience density line item collapses, and headcount is the metric institutional accounting systems measure.

Recent capital-response patterns illustrate the persistence of the substitutability assumption in practice. In May 2026, Hartford HealthCare announced a more than $1 billion campus overhaul targeting emergency department boarding following a 2025 average ED wait time of 8.9 hours at Hartford Hospital, the longest in Connecticut and among the longest nationally (Krechevsky, 2026). The response centers on a $950 million, 14-story inpatient tower with 216 additional patient beds, three procedural floors, a 1,650-car parking garage, and a 500-seat conference center. Workforce planning enters the announcement only as a single sentence indicating that the organization “plans to hire additional clinical and administrative workers to support the expansion.” Composition is unaddressed. The implicit model is that 216 new beds will be staffed by nurses interchangeable with any others, against composition mathematics that have already collapsed and that the present analysis documents. Capital responses of this scale, aimed at the throughput consequences of workforce-composition compression without engaging the composition variable itself, are the operational expression of the substitutability assumption Aiken et al. critiqued at the educational dimension and that this paper extends to the tenure dimension.

The pattern is not regional. In February 2026, Jefferson Health announced a $30 million campaign to expand the Abington Hospital emergency department from 80,000 to 100,000 annual visits, while in the same announcement noting that the hospital’s inpatient behavioral health unit had been closed the prior year to accommodate additional ED patients (Brubaker, 2026). The sequence makes the mechanism visible: contracting behavioral health capacity routes patients to the ED, which requires capital expansion, which displaces behavioral health capacity, which routes more patients to the ED.

A further asymmetry is worth naming, if only to set it aside. A bed tower is a capital asset that expands billable capacity and is financed as an investment; at Hartford, through roughly $850 million in bonds (Krechevsky, 2026). Competitive compensation to retain experienced clinicians is an operating expense that creates no asset and expands no billable capacity, even where, on the mechanism proposed in Section 4, it would reduce the throughput pressure the capital is built to relieve. Under fee-for-service reimbursement these are not weighted equally: capacity bills, retention costs. One would prefer to assume the asymmetry plays no part in the preference for building beds over retaining the clinicians who would reduce the need for them, and that the choice reflects only the measurement blind spot described above, and not an incentive. The present analysis cannot adjudicate which it is, and, indeed, it need not: the blind spot and the reimbursement structure point in the same direction.

Current hospital accounting and executive incentive structures may undervalue experience retention because short-term labor savings are visible immediately, whereas the downstream costs of experience loss emerge later as staffing instability, orientation burden, and reduced unit resilience. A pension contribution savings appears in a 2024 budget; the resulting departure of senior nurses retained by that contribution appears in a 2026 staffing event. Executive compensation horizons do not extend to the operational consequences of decisions made during executive tenure. This mismatch is reinforced by survey evidence from clinicians themselves: in a 2021 multi-site study of physicians and nurses across 60 U.S. Magnet hospitals, both groups ranked management interventions to improve staffing, work environments, and clinician control over workload as substantially more important to their well-being than wellness programs or resilience training (Aiken et al., 2023). The interventions hospital systems most commonly implement are not the interventions the workforce identifies as protective. Effective intervention requires either restructuring executive incentive horizons or imposing external constraints that make long-horizon experience-density costs visible in short-horizon decision frames.

## 8. What "acting now" looks like

The framing in Section 1 reshapes what counts as relevant policy. These are not narrowly nursing-workforce recommendations. They are healthcare-system-resilience recommendations that operate through nursing labor.

### Federal level

Expansion of Title VIII Nursing Workforce Development funding, with carve-outs for emergency nursing residency programs structured similarly to physician residencies, i.e., paid post-licensure structured training in the ED setting, replacing the current pattern of new-graduate ED hire with informal preceptor support. Expansion of HRSA Nurse Corps loan repayment programs targeted at retention in critical specialty settings, with particular attention to facilities serving populations that rely on the ED for residual access. Continuation of the NSSRN cycle on schedule with full methodological transparency.

### State level

California-model nurse-to-patient ratio legislation, with explicit acknowledgment that the cost-containment function of such legislation operates by making turnover costs visible in operating margins. Pension-contribution mandates analogous to public-employee pension protections, recognizing that hospitals serving residual-access populations are functionally performing a public service whose labor force warrants public-service-grade protections. State-board-of-nursing data reporting requirements that mirror NSSRN cell-level breakdowns, providing leading-indicator data when federal data is delayed or compromised.

Workforce investment alone, however, addresses only the supply side of the residual-access function the ED has come to perform. The underlying demand is generated by the absence of alternative access infrastructure for the populations described in Section 1, and remediation of workforce conditions inside the institution does not by itself reduce the volume of need being routed to it. A whole-systems response requires parallel investment in non-ED access modalities, of which three are particularly relevant to the residual-access function this paper has described: primary care access expansion for the populations using the ED for chronic-disease management and preventive care; community-based behavioral health and crisis services, including continued 988 implementation, mobile crisis response, and crisis stabilization unit capacity, for the populations using the ED for psychiatric crisis intervention; and community paramedicine for the populations whose presenting needs can be addressed safely in pre-hospital or community settings without ED transport.

Of these, community paramedicine warrants particular attention as a model whose international evidence base documents measurable ED-utilization reduction without compromising clinical outcomes (Hoyle et al., 2012; Swain et al., 2012; Lurie et al., 2023; Spelten et al., 2024), but whose U.S. implementation remains structurally constrained by regulatory fragmentation across state scope-of-practice frameworks (Glenn et al., 2018). U.S. implementation has been limited to demonstration projects and waiver-dependent pilots, with permanent CMS reimbursement pathways the principal structural barrier to scaling. The paramedic workforce faces compression dynamics broadly parallel to those documented here for emergency nursing, and the structural transferability of international community-paramedicine models to the U.S. context is the subject of forthcoming comparative work by the authors.

### Most fundamentally

institutional financial reporting that includes experience density as a measured operational metric next to headcount, so that the variable determining unit function becomes visible to the decision-makers whose decisions determine it. The clinicians themselves are clear about which interventions matter. In the largest recent multi-site survey of U.S. hospital clinicians, 87% of nurses and 45% of physicians ranked nurse staffing improvements as the highest-priority intervention for their well-being, while clinician wellness and resilience programs ranked lowest (Aiken et al., 2023). Institutional response should match clinician-identified priority rather than the lower-cost interventions that have predominated in hospital response to date.

## 9. Limitations

The 2018 and 2022 NSSRN cycles use slightly different sampling frames and questionnaire structures, which affect some cross-year comparisons. The 2018 cycle does not include the discrete EMT/paramedic prior-employment variable, preventing clean cross-year testing of the EMS pipeline. The 2022 cycle codes the teaching/precepting/orienting variable in coarser increments than 2018, restricting cross-year comparison to the binary "any involvement" construct. The Norful leaving-reasons proxy for burnout is operationally identical across cycles but conceptually different from the 2022 direct-question item; we use the proxy for cross-year comparison and the direct item only for 2022 sensitivity analysis.

Response rates declined materially between cycles, from 50.1% in 2018 to 40.6% in 2022. If respondents experiencing higher burnout were less likely to respond in 2022 (a fairly plausible direction for non-response bias in a survey burdening a workforce population in active distress) our cross-cycle burnout shift estimates are *conservative* relative to the true population shift. The 2026 cycle’s OMB submission explicitly cites lower response rates as a driver of increased per-respondent survey costs (OMB Supporting Statement A, 2025), suggesting the response rate trajectory continues; this strengthens rather than weakens the relevance of conservative cross-cycle estimates in identifying workforce composition change.

Publication analyses used replicate-weight variance estimation consistent with NSSRN technical documentation; preliminary WR analyses produced substantively similar point estimates and conclusions.

The residual-access framing in Section 1 is supported by published literature on ED utilization patterns; this paper documents workforce conditions inside the institution bearing that function rather than independently measuring the function. The defensive-utilization mechanism named in Section 4 is a hypothesis whose workforce-composition prerequisites are documented here but whose empirical confirmation requires linkage to operational data this paper does not access. Experience density is introduced as a construct in this paper rather than as an established literature variable; its operationalization here (years since first RN licensure plus tenure distribution of nurses with knowledge-transfer involvement) is one of several possible operationalizations and warrants further methodological development.

Years since first RN licensure tracks accumulated nursing experience across settings rather than ED-specific exposure. This is consistent with the experience-density construct as developed here (pattern recognition for clinical presentations, awareness of failure modes, capacity to transmit both to novice practitioners), which is not setting-bounded: senior nurses entering the ED from outpatient practice, surgical services, infusion, school nursing, or other contexts bring transferable clinical judgment that the licensure-tenure proxy correctly captures. The traveler-adjusted sensitivity reported in Section 3.3 addresses the related concern that licensure tenure may not adequately capture short-position-tenure exposure by adjusting for a behavioral marker of contract employment; the ED main effect was robust to that adjustment. The proxy does not, however, disaggregate the composition of late-career nurses’ tenure between ED and non-ED settings, and to the extent future work concerns itself with cross-setting career trajectories specifically, the public-use NSSRN files do not currently permit clean disaggregation.

The 2008 NSSRN cycle, presented in Section 3.1 as a structural baseline, was conducted under a different sampling design (independent state-stratified sampling, replacing the nested alpha-segment design used in 1977–2004) and introduced regression-based imputation for item nonresponse and differential nonresponse adjustment by age group. HRSA *explicitly* cautions against direct comparison of 2008 age-correlated estimates to prior cycles. The 2008→2018 transition was a more substantial redesign than 2018→2022, involving revised questionnaire content, a Census PVS-based sampling frame, and web-first instrumentation. We treat 2008 as a structural baseline reference rather than as a third co-equal analytic cycle: cross-cycle interpretation of 2008-to-2018 differences should acknowledge that observed change may reflect both genuine workforce dynamics and instrument-design discontinuity. Variance estimation for 2008 used standard jackknife (JK1) with 100 replicate weights and scale (R−1)/R. ED identification in 2008 used a clinical-content-based definition (Q29A_5 level of care, Q29C_7 specialty) because the 2008 employment setting variable does not include a discrete emergency department code; this is a methodologically distinct identification strategy from the current-position definition used for 2018 and 2022, reported here as a sensitivity-of-claim limitation rather than as a fully harmonized cross-cycle estimator. The 2008→2018 window is uninterrupted by intermediate NSSRN RN cycles: HRSA did not field a 2012 RN NSSRN (the 2012 National Sample Survey of Nurse Practitioners covered NPs only), and no 2014 or 2016 RN cycle was fielded. The decade between 2008 and 2018 is the longest unsurveyed gap in NSSRN’s history, during which the workforce changes documented here accumulated without federal cross-sectional measurement.

## Declarations

### Ethics approval

This study analyzed publicly available, fully de-identified secondary data from the NSSRN public-use files. It does not involve human-subjects research as defined under 45 CFR 46 and therefore did not require institutional review board review or approval.

### Data availability

The data analyzed in this study are the 2008, 2018, and 2022 National Sample Survey of Registered Nurses (NSSRN) public-use files, which are publicly available from the Health Resources and Services Administration (HRSA) at https://bhw.hrsa.gov/data-research/access-data-tools/national-sample-survey-registered-nurses.

### Code availability

Analysis code, including replicate-weight construction, domain estimation, and the regression specifications reported here, is available at https://doi.org/10.5281/zenodo.20576251.

